# Boosting the power of rare variant association studies by imputation using large-scale sequencing population

**DOI:** 10.1101/2023.10.28.23297722

**Authors:** Jinglan Dai, Yixin Zhang, Zaiming Li, Hongru Li, Sha Du, Dongfang You, Ruyang Zhang, Yang Zhao, Zhonghua Liu, David C. Christiani, Feng Chen, Sipeng Shen

**Affiliations:** Department of Biostatistics, Center for Global Health, School of Public Health, Nanjing Medical University, Nanjing 211166, China; Jiangsu Key Lab of Cancer Biomarkers, Prevention and Treatment, Jiangsu Collaborative Innovation Center for Cancer Personalized Medicine, Nanjing Medical University, 211166, Nanjing, China; China International Cooperation Center of Environment and Human Health, Nanjing Medical University; Key Laboratory of Biomedical Big Data of Nanjing Medical University, Nanjing 211166, China; Department of Biostatistics, Columbia University, New York, NY, USA; Department of Environmental Health, Harvard T.H. Chan School of Public Health, Harvard University, Boston, MA 02115, USA; Pulmonary and Critical Care Division, Massachusetts General Hospital, Department of Medicine, Harvard Medical School, Boston, MA 02114, USA

**Keywords:** rare variant, genotype imputation, whole-genome sequencing, association analysis

## Abstract

Rare variants can explain part of the heritability of complex traits that are ignored by conventional GWASs. The emergence of large-scale population sequencing data provides opportunities to study rare variants. However, few studies systematically evaluate the extent to which imputation using sequencing data can improve the power of rare variant association studies. Using whole genome sequencing (WGS) data (n = 150,119) as the ground truth, we described the landscape and evaluated the consistency of rare variants in SNP array (n = 488,377) imputed from TOPMed or HRC+UK10K in the UK Biobank, respectively. The TOPMed imputation covered more rare variants, and its imputation quality could reach 0.5 for even extremely rare variants. TOPMed-imputed data was closer to WGS in all MAC intervals for three ethnicities (average Cramer’s V>0.75). Furthermore, association tests were performed on 30 quantitative and 15 binary traits. Compared to WGS data, the identified rare variants in TOPMed-imputed data increased 27.71% for quantitative traits, while it could be improved by ∼10-fold for binary traits. In gene-based analysis, the signals in TOPMed-imputed data increased 111.45% for quantitative traits, and it identified 15 genes in total, while WGS only found 6 genes for binary traits. Finally, we harmonized SNP array and WGS data for lung cancer and epithelial ovarian cancer. More variants and genes could be identified than from WGS data alone, such as *BRCA1*, *BRCA2*, and *CHRNA5*. Our findings highlighted that incorporating rare variants imputed from large-scale sequencing populations could greatly boost the power of GWAS.

## Introduction

Conventional genome-wide association studies (GWASs) are well-powered to detect thousands of common variants associated with human traits and diseases [1–3]. However, GWASs underrepresent rare variants due to the limitations of SNP array [4]. Rare variants have larger effects and behave differently from common variants to explain a fraction of human traits or disease heritability [5, 6]. With the emergence of next-generation sequencing (NGS) [e.g., whole genome sequencing (WGS)] based on biobank-level populations, massive rare variants [minor allele frequency (MAF)<0.01] or ultra-rare variants (MAF<0.0001) could be accurately captured [7]. For example, several studies aimed at identifying novel germline variants in cancer or rare diseases have benefited from NGS [8, 9].

However, given that the application of deep WGS is limited by cost for large-scale populations [10], SNP array is a cost-effective and major approach for human genomics exploration thus far, such as UCLA ATLAS [11], China Kadoorie Biobank [12], FinnGen [13], and various caseLcontrol GWASs [14]. Even if there are high-quality population-based sequencing data, the incidence number of complex disease cases is often low and the power for identifying rare variants is insufficient compared to caseLcontrol studies based on SNP array. For example, the UK Biobank (UKB) has whole-exome sequencing (WES) data with a large sample size (n ≈ 450,000) [15, 16], but most cancer cases have a sample size of less than 10,000. Nevertheless, unlike sequencing data, SNP array-based genotyping can only represent a fraction of the genetic variation in the genome. Thus, it is a more appropriate method to power association analysis to detect rare variants that were not genotyped to the SNP array by imputing from the external high-quality large-scale sequencing panels [10, 17], such as the UK10K, Haplotype Reference Consortium (HRC) and Trans-Omics for Precision Medicine (TOPMed). In TOPMed, 290 million (∼97%) variants had an MAF <1%, which might be strong candidates for association analyses [18, 19]. However, few studies have focused on evaluating the accuracy of rare variant imputation based on sequencing data panel and the extent to which it can enhance the power of rare variant exploration.

Here, we present a comparative study that leverages the WGS data of 150,119 individuals from the UKB as the ground truth to evaluate the coverage and accuracy of rare variant imputation using HRC+UK10K and TOPMed. We investigated the association tests of WGS data and imputed variants with 30 biochemistry biomarkers and 15 complex diseases, respectively. Finally, we harmonized WGS data and diverse SNP array data to reveal the rare variant signals in lung cancer and epithelial ovarian cancer.

## Results

### Landscape of rare variant imputation results

We analyzed WGS data from 150,119 individuals and genotype imputation data generated from the HRC+UK10K and TOPMed reference panels (n=488,377) from the UKB, respectively (Figure 1). The intersection of variants between WGS and imputed data is shown in Figure 2A. TOPMed imputed a substantially larger number of genetic variants that could detect approximately 26% single nucleotide variants (SNVs) of WGS data, which was higher than the 10.7% detected in HRC+UK10K. The TOPMed-imputed data could detect 22 million singleton and doubleton variants, which was far more than HRC+UK10K-imputed data but was still fewer than 333 million variants in WGS data. For ultra-rare variants, the TOPMed-imputed variants could reach 66.6% of WGS and were ∼3.4-fold of that in HRC+UK10K. For rare and common variants, the number of imputed variants was close to WGS (Figure 2B and Table S1). For protein-coding variants, TOPMed-imputed data detected 44,440 loss-of-function (LoF) variants, 509,759 synonymous variants and 880,589 missense variants, which was at least four-fold greater than that in the HRC+UK10K imputation (Figure 2B).

**Figure 1.**
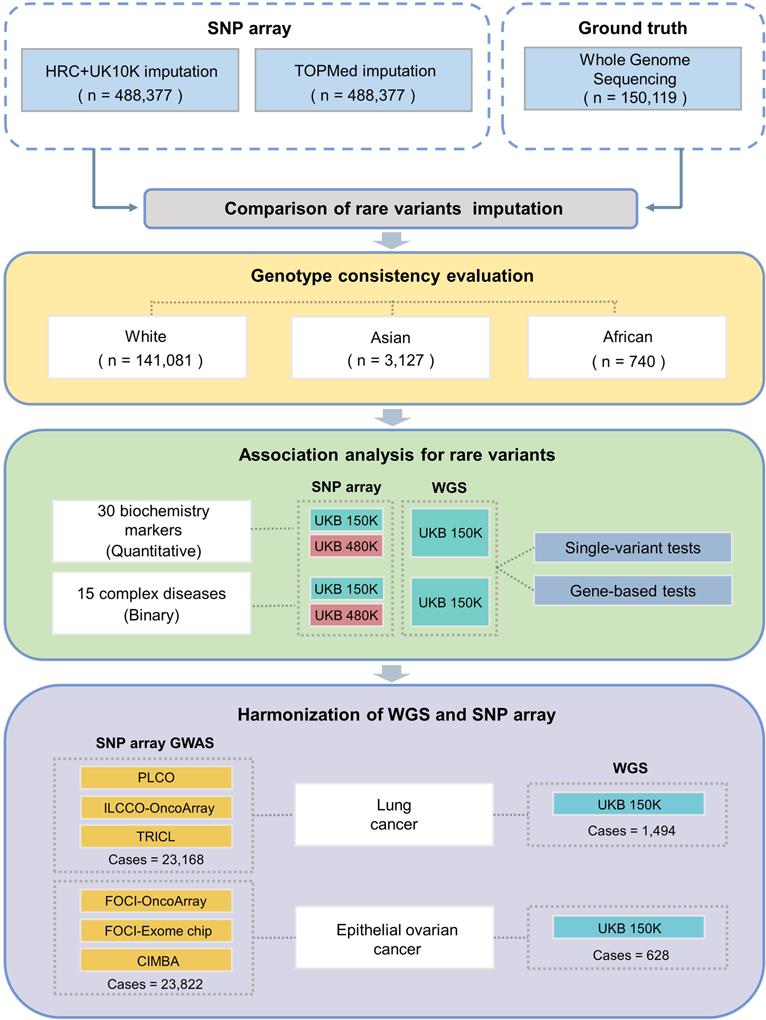
Study workflow. The schematic of the analytical pipeline summarizes the main steps for conducting a comparative study using WGS as the ground truth. We first compared the rare variant imputation for two imputed data (TOPMed-imputed and HRC+UK10K imputed) and WGS data. Then genotype consistency evaluation was investigated on three ethnicities. The rare variant association analysis of 45 traits was carried out after above evaluation in different data. Finally, we harmonized the WGS and SNP-array data for the two cancers.

**Figure 2.**
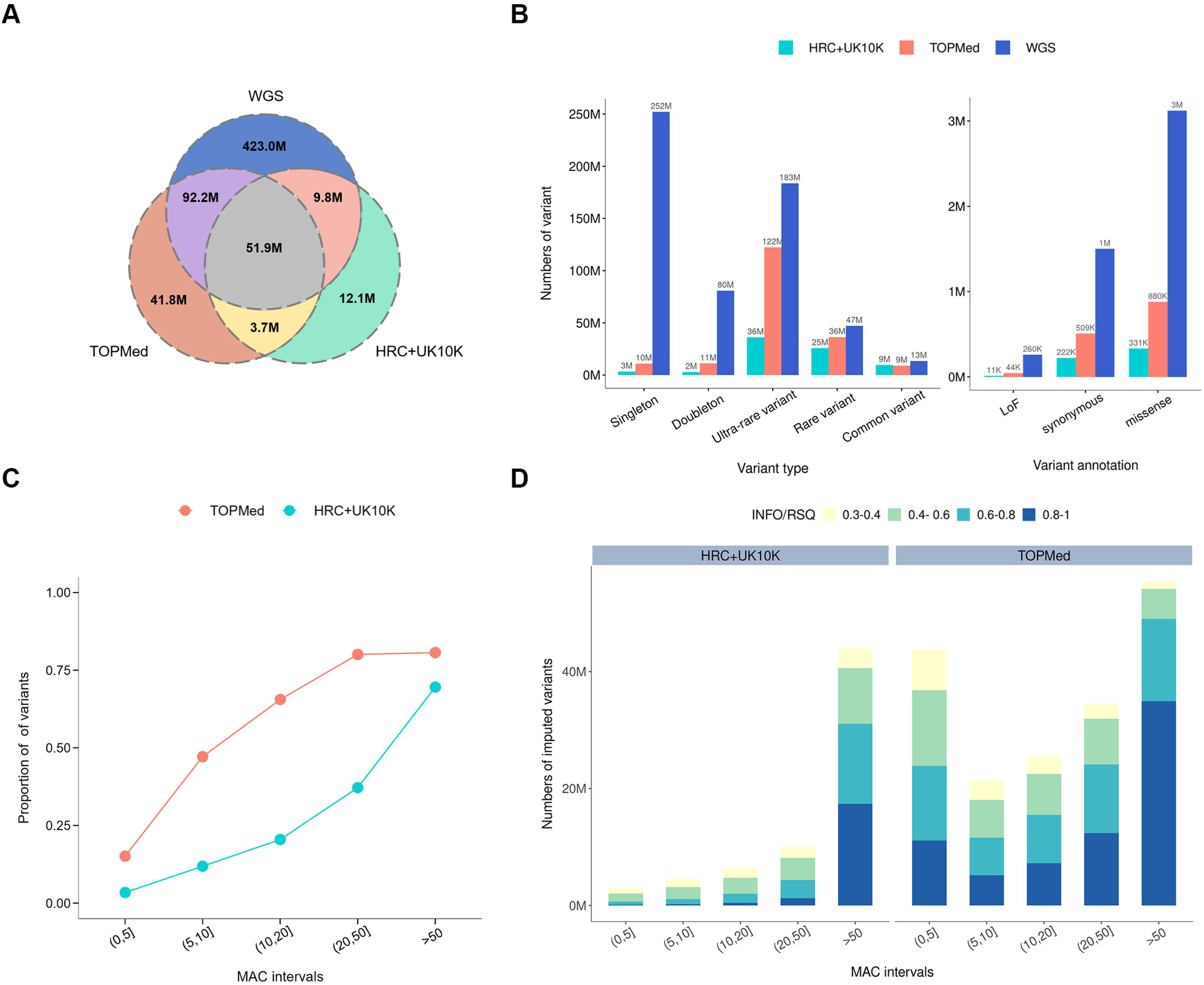
Landscape of rare variant imputation results. (A) Venn diagram showing the intersection of variants between TOPMed-imputed data, HRC+UK10K-imputed data and WGS data. (B) Bar chart showing the number of five distinct variant types for different data (left). **Variant type descriptio: Singleton: The minor allele occurs only once in the entire sample Doubleton: The minor allele occurs twice in the entire sample Ultra-rare variant: MAF<0.0001 except singleton and doubleton Rare variant: MAF in 0.0001-0.01 Common variant: MAF>0.01 Bar chart showing the number of three distinct variant annotations for different data (right). (C) Line chart of variant coverage proportions in different imputed data. The x-axis represents the MAC intervals, and the y-axis represents the proportion of variants that imputed data could detect related to WGS data. (D) The number of variants imputed by different panels and their distribution across INFO/RSQ bins.

To deeply understand the performance of different imputed data compared to WGS, all variants were divided into five intervals according to the minor allele count (MAC). As expected, we observed an increase in the fraction of variants as MAC increased. When MAC∈(10,20], TOPMed-imputed data had already detected ∼65% variants of WGS data, HRC+UK10K-imputed data could only detect ∼20% (Figure 2C and Table S2).

For the imputed data, post-imputation quality control (QC) is generally performed based on INFO/RSQ filtering. The number of different INFO/RSQ variants in the imputed data is presented in Figure 2D. TOPMed-imputed data achieved substantially higher coverage of rare variants than HRC+UK10K, and as the MAC increased, the proportion of high-quality TOPMed-imputed variants with higher INFO/RSQ increased. Even for extremely rare variants (MAC∈(0,5)), the average INFO/RSQ of imputed data could also reach 0.5 (Table S3).

### Evaluation of genotype consistency between WGS and imputed data

We used WGS data as the ground truth to conduct correlation analysis on the two imputed datasets to evaluate the consistency of rare variants in three ethnicities (White, Asian and African) (Table S4). TOPMed had a better imputation performance that was closer to WGS than HRC+UK10K in all MAC intervals, and the average Cramer’s V imputed by TOPMed was above 0.75 in three ethnicities (Figure 3A and Table S5). Even for MAC∈(0,5), the Cramer’s V of TOPMed-imputed data exceeded 0.6 in all ethnicities. The difference in Cramer’s V of various intervals between TOPMed-imputed data and HRC+UK10K-imputed data reached a maximum in extremely rare variants (MAC (0, 5)), which was 0.33 for White, 0.25 for Asian and 0.17 for African (Figure 3A and Table S5).

**Figure 3.**
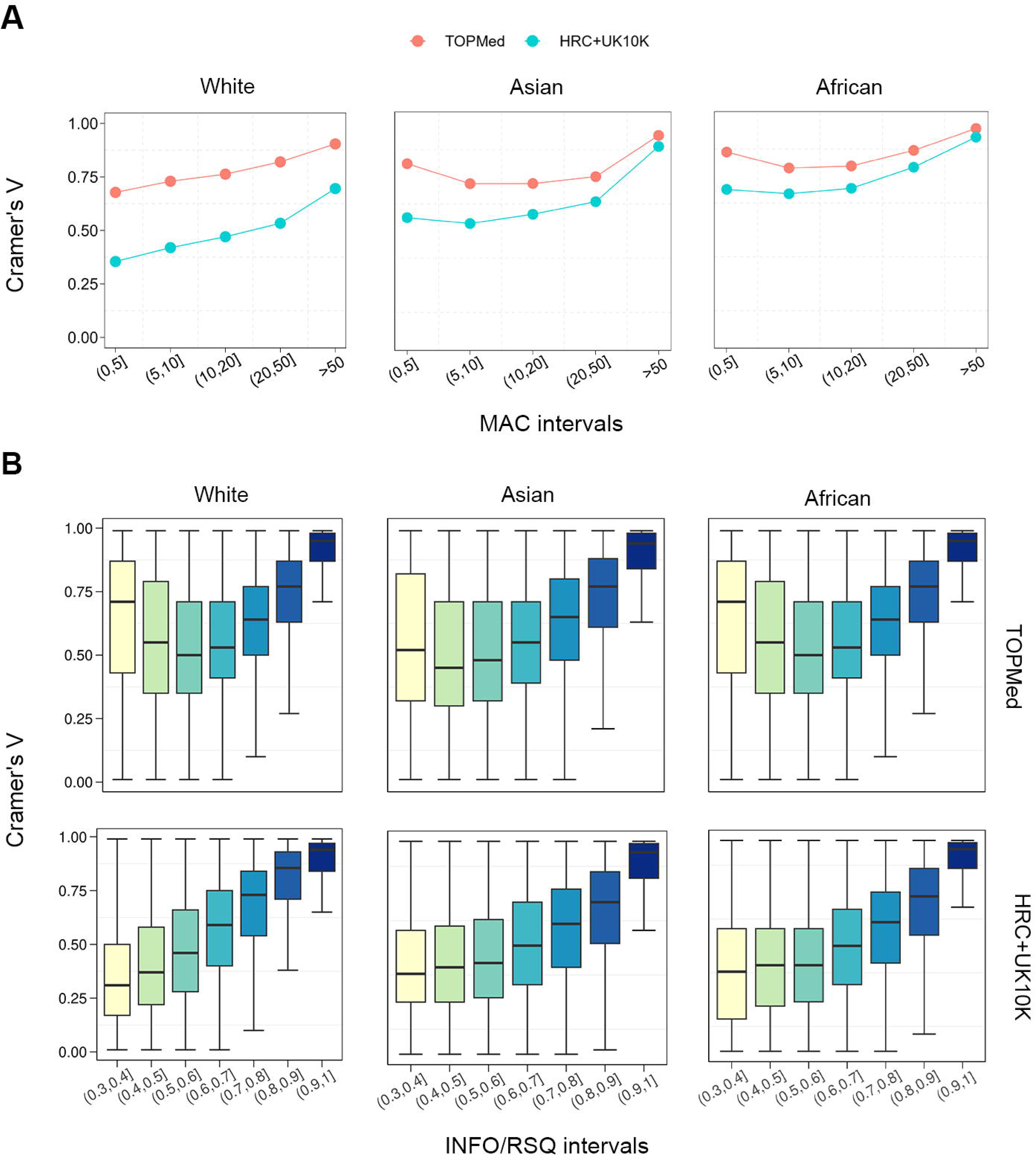
Consistency evaluation of the imputed datasets in different ethnicities. (A) Correlation of three ethnicities between imputed data and WGS data in different MAC intervals. (B) Box plot of the relationship between the INFO/RSQ of imputed data and the Cramer’s V.

We also described the relationship between INFO/RSQ and Cramer’s V (Figure 3B). With the increase in INFO/RSQ bins, Cramer’s V also increased in all three ethnicities, indicating that the variants with high imputation quality could be closer to the WGS data to some extent. Moreover, for the same INFO/RSQ bin, using different reference panels would obtain different Cramer’s V, and the consistency of TOPMed-imputed data was higher than that of HRC+UK10K in each bin. Even when the imputation quality was not particularly good (INFO/RSQ ∈ (0.3,0.4]), the TOPMed-imputed data had an advantage over that from HRC+UK10K which showed a stable consistency (all Cramer’s V>0.5 in three ethnicities) (Table S6). Therefore, the rare variants imputed from TOPMed were applicable for subsequent association analysis.

### Rare variant association analysis for quantitative biochemistry biomarkers

We performed association tests between rare variants (MAF<0.01) and 30 biochemistry biomarkers in WGS (n=150 k) and imputed data (n=150 k and 480 k) of European descent. In the single-variant tests of 16 traits, compared to WGS data, the two imputed data (n=480 k) could detect more significant rare variants; HRC+UK10K-imputed data could improve 4.7% while the TOPMed-imputed data could improve 27.71% (Figure 4A-4B). However, the number of significant rare variants found in WGS data was more than that of imputed data in 12 traits, which might be due to the extremely large number of sequencing sites in WGS. Meanwhile, we noticed that the imputed data of n=150 k had a weaker ability to detect rare variants than WGS (Figure S2). On average, the number of significant rare variants in the TOPMed-imputed data was 661 and in the HRC+UK10K-imputed data was 450, which were less than 2,479 in WGS data (Table S7).

**Figure 4.**
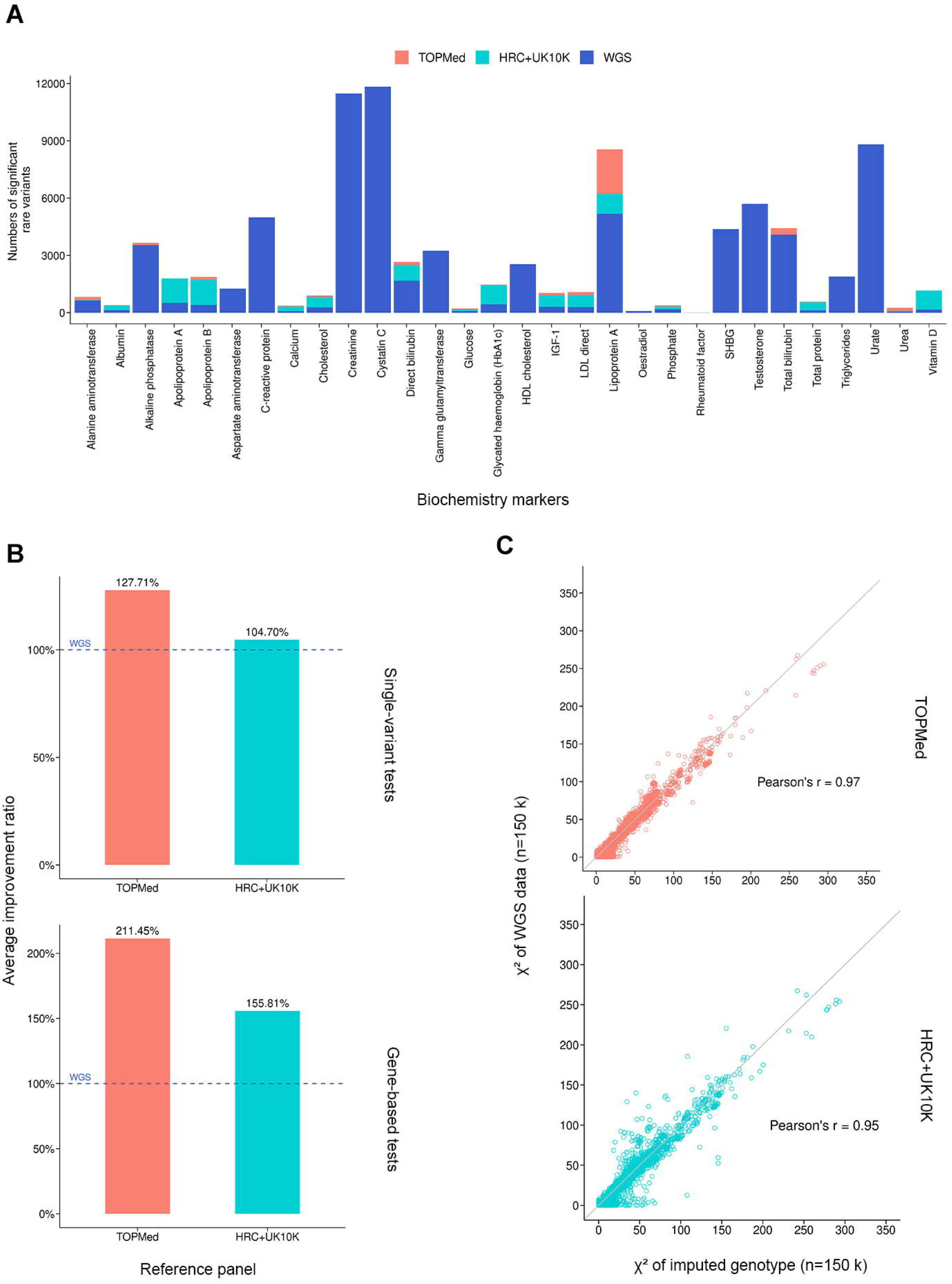
Rare variant association analysis results for biochemistry biomarkers. (A) The number of additional significant rare variants (MAF<0.01) found in the imputed data compared to the WGS data for 30 quantitative traits. (B) The average improvement ratio compared to WGS data for single-variant tests and gene-based tests. (C) Pairwise Pearson correlations between chi-square statistics produced by association tests using imputed data (n=150 k) and WGS data respectively.

We also evaluated the gene-based tests results to capture the effects of rare variants. On average, 22 genes were found in the TOPMed-imputed data and 18 genes were found in the HRC+UK10K-imputed data (n=480 k), while the WGS data identified only 7 genes for all traits (Table S8). The average improvement ratio of the HRC+UK10K-imputed data was 55.81%, and that of the TOPMed-imputed data reached 111.45% (Figure 4B). When comparing the WGS data with the imputed data of n=150 k, the number of significant genes in the TOPMed-imputed data decreased by 27.10%, while that in the HRC+UK10K-imputed data decreased by 33.66% (Figure S2).

Based on these association tests of the significant variants, we depicted the relationship between the chi-square statistics of the association analysis in WGS data and two imputed data (n=150 k). The test statistics in different imputed data both presented a strong relationship with that in WGS data, which meant their association test statistics were highly correlated (on average, Pearson’s *r* =L0.97 for TOPMed-imputed data and 0.95 for HRC+UK10K-imputed data) (Figure 4C and Table S9). Therefore, the rare variant-trait associations using imputed genotypes were robust.

### Rare variant association analysis for complex diseases

We also conducted single-variant tests and gene-based tests for 15 complex diseases, including ten chronic diseases and five cancers (Table S10). In single-variant tests, the number of significant rare variants in the imputed data (n=480 k) was approximately 10-fold that in WGS data, owing to its larger sample sizes with sufficient disease cases (Figure S1 and Table S13). On average, 22 significant rare variants were identified in TOPMed-imputed data, and 24 were identified in HRC+UK10K-imputed data. However, only 2 variants could be found in the WGS data on average (Figure 5 and Table S11). We also found that for the imputed data with a sample size of 150 k, its ability to detect rare variants was a slightly weaker than that of WGS (Figure S3). Therefore, it could be expected that using large-scale SNP array-based data imputed from sequencing panels would greatly improve the ability of rare variant findings compared to WGS data.

**Figure 5.**
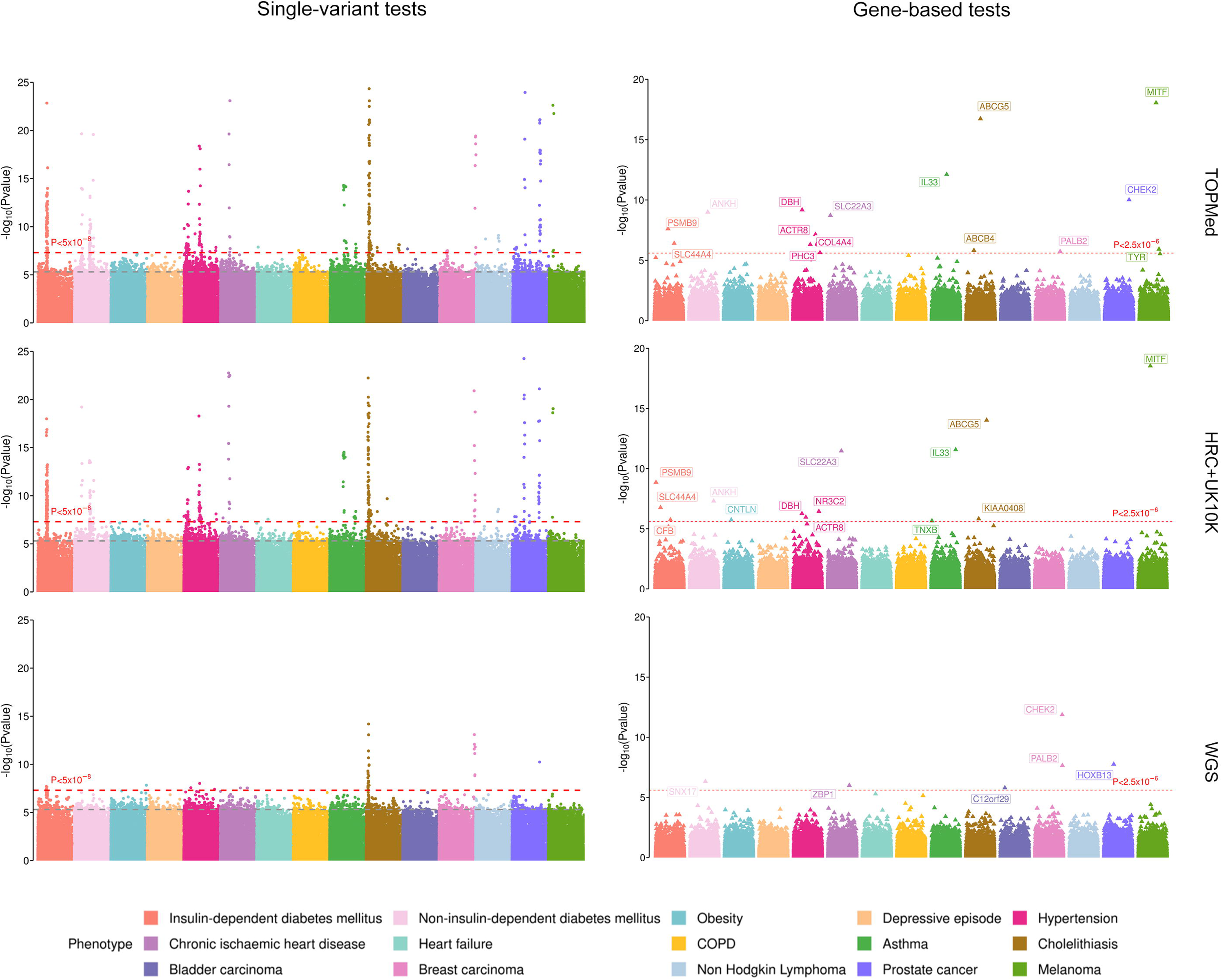
Rare variant association analysis results for complex diseases. Multiple-trait Manhattan plot of single-variant tests and gene-based tests for 15 diseases in different data. The x-axis labels each disease, and the y-axis shows -log_10_*P*. The red dotted line represents the significance filtering threshold of the *P* value (*P* < 5.10^-8^ for single-variant tests, *P* < 2.5.10^-6^ for gene-based tests), and the gray dotted line represents the suggestive filtering threshold of the *P* value (*P* < 5.10^-6^ for single-variant tests)

In the gene-based tests, the number of significant genes in the imputed data was higher than that in the WGS data, and the performance of the TOPMed-imputed data was better. HRC+UK10K-imputed data could find 14 significant genes in total, and the TOPMed-imputed data could find 15 genes, while the WGS data only found six genes owing to low cases (Figure 5 and Table S12, S14). In addition, we also noticed that some classical genes identified in the two imputed datasets were roughly consistent but undetected in WGS. For example, *IL33* has been certified to be associated with asthma and a rare mutation in *IL33* could decrease the risk of asthma [20, 21]. For prostate cancer, the gene *CHEK2* which was found only in TOPMed-imputed data has been confirmed to be a risk gene containing rare variants [22]. Most genes found in the TOPMed-imputed data had supporting association or biological evidence from previous studies (Table S21).

Based on the results above, we concluded that incorporating SNP array imputed from large-scale sequencing populations could enhance the ability to detect rare variant-trait associations.

### Powering the rare variant hits by harmonizing WGS and SNP array

Furthermore, we leveraged UKB WGS and several SNP array data imputed by the TOPMed reference panel to perform association tests for lung cancer (LC) and epithelial ovarian cancer (EOC) (Figure 6A, 6D). Compared with limited cases in WGS (n_LC_ = 1,494 ; n_EOC_ = 628), the SNP array could provide sufficient cases (n_LC_ = 23,168; n_EOC_ = 23,822) that were 16-fold and 38-fold for LC and EOC respectively. In single-variant tests, no rare variants could pass the genome-wide significance level in WGS. However, when combined with SNP-array data, we detected 12 SNVs and 22 SNVs, mapping to six and four independent genetic regions, respectively (Figure 6B, 6E, Table S17-S18). In gene-based tests, no genes could pass *P*< 2.5×10^-6^ in WGS. In the meta-analysis, we identified *BRCA2* and *CHRNA5* in lung cancer, while *BRCA2* and *BRCA1* were significant in EOC (Figure 6B, 6E, Table S19-S20). Using different *P* value thresholds, the WGS+SNP-array strategy could detect more variants, especially under stringent thresholds (e.g., 10^-7^ in single-variant and 10^-5^ in gene-based) (Figure 6C, 6F).

**Figure 6.**
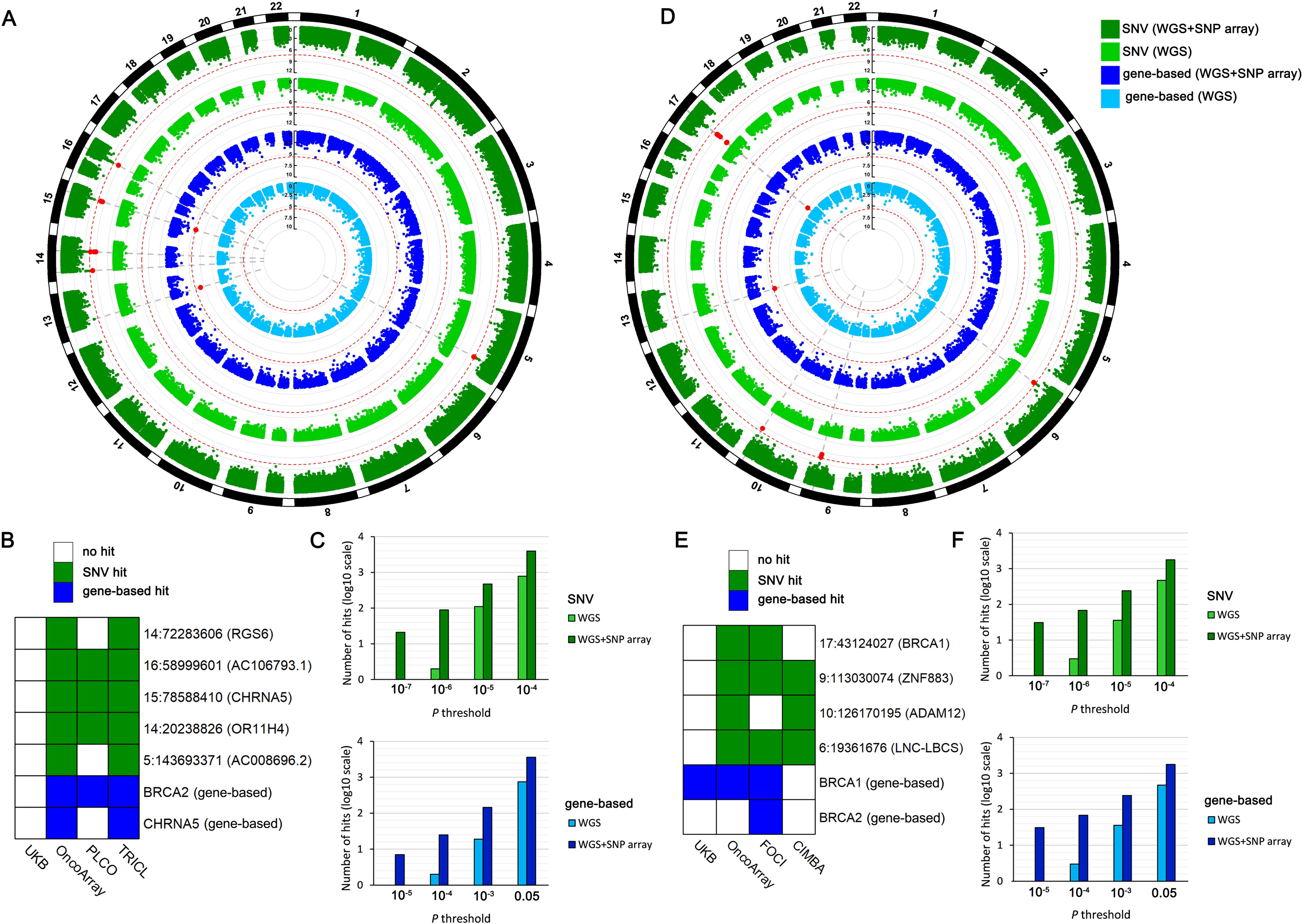
Harmonizing WGS and SNP array to perform association tests for lung cancer (left panel) and epithelial ovarian cancer (right panel) (A-C) Results of lung cancer. (D-F) Results of epithelial ovarian cancer. (A, D) Circos plots of the single-variant and gene-based association results using UKB WGS data or WGS+SNP array data. (B, E) Rare variants and genes identified in each dataset. The blocks are marked if the *P* values reach nominal significance (*P*<0.05). (C, F) Comparison of the identified signals in the single-variant and gene-based association tests under different *P* value thresholds.

## Discussion

It is a common practice to directly apply sequencing data or imputed genotype data to GWAS. Limited by the cost of NGS technology, biobank-level sequencing data are still in the minority [23], resulting in the difficulty of using rare variant analysis which provides important genomic architectures. Large-scale GWASs tend to use imputed genotype data, but few studies systematically compare the power of imputed data for rare variants. Therefore, evaluating the consistency and association power performance of rare variants imputed from sequencing data can provide better guidance and reference for rare variant exploration in complex traits.

In this study, we comprehensively conducted a series of comparative analyses for rare variant imputation using SNP array data from the UKB based on TOPMed or HRC+UK10K against the WGS data of 150,119 individuals as the ground truth. We described the rare variant imputation results, and further carried out the correlation analysis in three ethnicities (White, Asian, and African) to evaluate how close rare variant imputation could be to WGS data. We further investigated association analysis on 30 biochemistry markers and 15 complex diseases to explain the ability of imputed rare variants to improve the association analysis at both single-variant and gene-based levels.

Our summarized description shows that the TOPMed panel could impute more high-quality rare variants than HRC+UK10K. Although the imputed variants were far inferior to WGS in singleton and doubleton variants, the total number of rare variant imputed by TOPMed also had a certain scale. Even for the extremely rare variants (MAC<5), the average imputation quality of TOPMed-imputed variants was acceptable (INFO/RSQ≥0.5). Although not all rare variants can be estimated reliably, they could provide a crucial supplement in addition to WGS.

For the three ethnicities, we observed that rare variants imputed from TOPMed were closer to WGS than those imputed from HRC+UK10K in each MAC interval. In Europeans, the consistency of rare variants with MAC≤20 in HRC+UK10K-imputed data is poor, which is not recommended for subsequent application. In addition, the overall correlation between TOPMed-imputed data and WGS was slightly stronger in Africans than in the other two ethnicities, which may be due to the low sample size of Africans (n<1,000) in WGS, and unbalanced ethnic sample sizes affect the stability of Cramer’s V. Nevertheless, the TOPMed-imputed data still had a stable imputation performance that was closer to WGS data, indicating its reliability to be applied in rare variant association studies.

In the association analysis of quantitative and binary traits, although the ability of both imputed data to identify significant rare variants was weaker than that of WGS data in the same population (n = 150k), we could still identify a few rare variants or gene sets. When the sample size of the imputed data increased to n = 480 k, the ability to identify significant rare variant could improve, but this improvement was slightly different in distinct types of traits. For quantitative traits, the power of WGS data for finding significant rare variants was basically sufficient, and the two imputed data had limited improvement compared to WGS data. However, for binary traits, the number of disease cases in WGS data was far from sufficient to ensure optimistic power, causing fewer significant rare variants to be found. To improve the power, external data with additional disease cases must be supplemented. Our study indeed showed that larger SNP array data imputed from sequencing data could greatly improve the ability to find significant rare variants.

Although the natural population cohorts are large with adequate sample sizes, the disease cases are generally insufficient, which is known as a caseLcontrol imbalance problem and leads to low statistical power to identify rare variants [24]. Here, we harmonize WGS population and multiple caseLcontrol design GWASs and successfully identify classical rare variants and gene sets in two cancers, including the well-known *BRCA2* [25], *BRCA1* [26], and *CHRNA5* [27], which could not be detected in UKB WGS. Therefore, integrating SNP-array data through imputation is of great significance for discovering rare variants.

There are several strengths in our study. First, we comprehensively described the landscape of the imputed rare variants from different sequencing panels in terms of variant amount, coverage, and imputed quality. Second, we leveraged WGS data as the ground truth to perform correlation analysis by different ethnicities, and analyzed their consistency with imputed rare variants. We demonstrated the feasibility of using imputed rare variants for association analysis. Third, we conducted various association tests on 45 traits of the UKB and harmonized six GWAS caseLcontrol datasets totally. Through both single-variant and gene-based tests, we quantitatively explained the improvement of SNP array-based data imputed from sequencing data in rare variant association studies.

The results we presented here also have some limitations. First, our genotype consistency evaluation was only conducted on the UKB SNP array (∼50,000 UK BiLEVE Axiom array and ∼450,000 UK Biobank Axiom array) that did not consider other array types, which might influence the results. Second, we currently impute the genotype in TOPMed’s imputation online server (97,256 reference samples) due to computing power limitations, but it is convenient and feasible for most genomic studies. Theoretically, the results of imputation in larger populations (e.g., UKB whole WGS population) should be more accurate [28], which needs further studies to confirm. Third, we attempted to integrate the SNP-array data and sequencing data in two exemplary cancers. The effect of integrating other various diseases in large-scale population cohorts (n≥100,000) needs further evaluation.

In conclusion, incorporating rare variants imputed from large-scale sequencing populations can greatly enhance the power of GWAS research. Our study shows that combining multi-source genomic data with a sufficient number of cases can accurately identify a wider range of rare variants, allowing us to take full advantage of different types of genetic data and to gain a deeper understanding of the causal links between rare genetic variants in human complex traits and diseases.

## Methods

### UK Biobank population and phenotypic data collection

The UK Biobank (UKB) is a population-based prospective cohort of individuals aged 40–69 years, enrolled between 2006 and 2010 [29]. The work described herein was approved by the UK Biobank under applications no. 92675 and 83445. All phenotypic data were accessed in July 2022.

Blood biochemistry data were collected from Category 17518. The UK Biobank embarked on a project to measure a wide range of biochemical markers in biological samples collected at baseline (2006-2010) in all 500,000 participants. All 30 biochemistry biomarkers were included as quantitative traits.

Health-related outcomes were ascertained via individual record linkage to national cancer and mortality registries and hospital in-patient encounters. Cancer diagnoses were coded by International Classification of Diseases version 10 (ICD-10) codes from data fields 41270 (Diagnoses - ICD10), 41202 (Diagnoses - main ICD10), and 40001 (primary cause of death: ICD10). Individuals with at least one recorded incident diagnosis were defined as cases. We included 15 common chronic diseases or cancers as binary traits, including insulin-dependent diabetes mellitus, non-insulin-dependent diabetes mellitus, obesity, depressive episode, hypertension, chronic ischemic heart disease, heart failure, COPD, asthma, cholelithiasis, bladder carcinoma, breast carcinoma, non-Hodgkin lymphoma, prostate cancer, and Melanoma.

### UK Biobank genomics data collection

We collected whole-genome sequencing (WGS) of 150,119 people in the UK Biobank (data field 23352) [30], which were sequenced to an average coverage of 32.5× (at least 23.5× per individual) using Illumina NovaSeq sequencing machines at deCODE Genetics (90,667 individuals) and the Wellcome Trust Sanger Institute (59,452 individuals). Sequence reads were mapped to the human reference genome GRCh38 using BWA. SNPs and short indels were jointly called over all individuals using GraphTyper (v2.7.1) [31], which provided more accurate genotype calls. This constitutes a set of high-quality variants, including 585,040,410 single-nucleotide variants (SNVs) and 58,707,036 indels.

We also collected imputed genotype data based on two mainstream reference panels. The first is the Haplotype Reference Consortium (HRC) and UK10K haplotype resource, which increases the number of testable variants over 100-fold to ∼96 million variants (data field 22828). The genetic data was imputed using two different reference panels. The HRC panel (64,976 haplotypes) was used wherever possible, but for SNPs not in that reference panel the UK10K + 1000 Genomes panel (12,570 haplotypes) was used [32]. The raw positions are in GRCh37 coordinates and then lifted over to GRCh38.

The second is imputation from genotype using the TOPMed R2 panel (97,256 deeply sequenced genomes), performed by the TOPMed Informatics Research Center [33]. After phasing the UK Biobank genetic data (carried out on 81 chromosomal chunks using Eagle v.2.4), the phased data were converted from GRCh37 to GRCh38 using LiftOver. Imputation was performed using Minimac4 v1.0.2. Imputation was performed in 1 Mb chunks and merged back together by chromosome.

### Quality control of imputed SNP array data

If an imputed variant on N samples has an imputation quality metric scored at *α*, it implies that the statistical power of association tests is approximately equivalent to *αN* perfectly observed genotype data [34]. To perform GWAS on the UK Biobank data with ∼480,000 samples, it is typical to use variants with imputation quality higher than 0.3, equivalent to ∼150,000 perfectly observed samples (WGS sample size). Thus, markers with poor imputation quality were not retained in the subsequent analysis (excluding the Minimac4 imputation quality metric RSQ < 0.3 or IMPUTE imputation quality metric INFO < 0.3).

### Genotype consistency evaluation

In two imputed datasets (TOPMed and HRC+UK10K) and WGS data, we divided the population into three ethnicities --- White, Asian, African based on ethnic data field 21000. To evaluate the consistency between the imputed genotype and WGS data, we matched the imputed data (n=480k) with the WGS data (n=150k) according to the individual ID of each ethnicity.

Then, we used PLINK2.0 to calculate the number of minor allele count (MAC) and minor allele frequency (MAF) of genetic variants in each ethnicity. Meanwhile, based on MAC and MAF, genetic variants were classified into five categories, namely (0,5], (5,10], (10,20], (20,50], >50 and MAF<0.01 (the interval was abbreviated as > 50 in the subsequent analysis). We chose Cramer’s V as the metric to evaluate the consistency of imputed variants and sequencing variants:

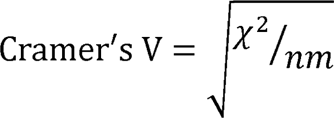

where χ^2^ represents the chi-square value of the current contingency table, n represents the sample size, and m represents the smaller value of the two degrees of freedom (r-1) or (c-1) of the two variables.

### Single-variant and gene-based association tests

Single-variant and gene-based association analyses were performed using REGENIE v3.2.6 [35]. It is a machine-learning based method to fit a whole-genome regression model for quantitative and binary phenotypes. Quantitative traits were rank-based inverse normal transformed. Saddlepoint approximation (SPA) was used to handle extreme caseLcontrol imbalance of binary traits. All association tests were performed in the population of European descent (ethnicity = “White”) only.

The single-variant association tests included rare variants with MAF < 0.01. The variants were functionally annotated using Variant Effect Predictor (VEP) software [36]. The genome-wide significant threshold of single-variant tests was set to *P* < 5× 10^-8^.

For gene-based analysis, we included rare and ultra-rare variants with MAF < 0.01. Three genetic models were considered: loss of function (LoF), LoF+missense and LoF+missense+synonymous. Of all the combinations, we reported the association results with the lowest *P* value to collectively capture a wide range of genetic architectures [5]. The genome-wide significance threshold of gene-based tests was set to *P* < 2.5×10^-6^.

In the association analyses of quantitative traits, we adjusted the covariates including age, sex, and the top ten principal components (PCs). In the association analyses of binary traits, we adjusted the covariates including age, sex (excluding sex-specific diseases), body mass index (BMI), smoking status (binary), drinking status (binary), and the top ten principal components (PCs).

### SNP array data collection for lung cancer and ovarian cancer

We also collected large-scale SNP array data of lung cancer and epithelial ovarian cancer to harmonize them with UKB WGS to improve the power.

For lung cancer, we investigated the association of imputed genetic variants with lung cancer in three additional cohorts from the Prostate, Lung, Colorectal, and Ovarian (PLCO) cancer screening trial [37], the International Lung Cancer OncoArray Consortium (ILCCO-OncoArray) [38], and the Transdisciplinary Research in Cancer of the Lung (TRICL) research team [39] (Table S15).

For ovarian cancer, we collected data in three additional cohorts from The Follow-up of Ovarian Cancer Genetic Association and Interaction Studies (FOCI)-OncoArray [40], FOCI-Exome Chip [41], and Consortium of Investigators of Modifiers of BRCA1/2 (CIMBA) [42] (Table S16).

Samples excluded were those who lacked disease status, were second-degree relatives or closer having identity by descent (IBD) > 0.2 or had low-quality DNA (call rate < 95%), or sex inconsistency, or were non-European. SNPs were removed if they met of the following criteria: (1) sex chromosome, (2) MAF < 0.05, (3) call rate < 95%, and (4) Hardy-Weinberg equilibrium (HWE) test *P* < 1.00×10^−7^ in controls or *P* < 1.00×10^−12^ in cases.

All genotype data were imputed on the TOPMed online imputation server. Poorly imputed SNVs with imputation quality score *R^2^* < 0.3 and SNVs on sex chromosomes were excluded from the analyses. The effect sizes and 95% confidence interval (CI) of genes were estimated by burden tests and then summarized by fix-effects meta-analysis.

## Declarations

### Consent for publication

All authors have reviewed and approved this manuscript.

### Data availability

UK Biobank data is available from: https://www.ukbiobank.ac.uk/.

ILCCO-Oncoarray data is available from: https://www.ncbi.nlm.nih.gov/projects/gap/cg-ibin/study.cgi?study_id=phs001273.v3.p2

TRICL data is available from: https://www.ncbi.nlm.nih.gov/projects/gap/cgi-bin/study.cgi?study_id=phs001681.v1.p1

PLCO data is available from: https://www.ncbi.nlm.nih.gov/projects/gap/cgi-bin/study.cgi-study_id=phs001286.v2.p2.

FOCI-Exome Chip data is available from: https://www.ncbi.nlm.nih.gov/projects/gap/cgi-bin/study.cgi?study_id=phs001131.v1.p1

FOCI-OncoArray Chip: data is available from: https://www.ncbi.nlm.nih.gov/projects/gap/cgi-bin/study.cgi?study_id=phs001882.v1.p1&phv=435464&phd=&pha=&pht=10492&phvf=&phdf=&phaf=&phtf=&dssp=1&consent=&temp=1

CIMBA: data is available from: https://www.ncbi.nlm.nih.gov/projects/gap/cgi-bin/study.cgi?study_id=phs001321.v1.p1

### Online resources

TOPMED imputation server: https://imputation.biodatacatalyst.nhlbi.nih.gov/. gnomAD: http://gnomad.broadinstitute.org/. dnSNP: https://www.ncbi.nlm.nih.gov/snp/.

### Code availability

The codes that support our findings are available from the corresponding author by a request.

### Author contributions

SS and FC contributed to the study design. SS and JD contributed to data collection. JD performed statistical analyses and interpretation. JD and SS drafted the manuscript. HL, SD, DY, YZ, RZ, ZL, and DC revised the final manuscript. All authors approved the final version of the manuscript.

## Supporting information

SuppFigures

SuppTables

## Acknowledgement

We want to acknowledge the participants and investigators of UK Biobank, TOPMed, PLCO, ILCCO-OncoArray, TRICL, FOCI, and CIMBA.

