## Supplementary material for "Boosting the power of rare variant association studies by imputation using large-scale sequencing population": SuppFigures

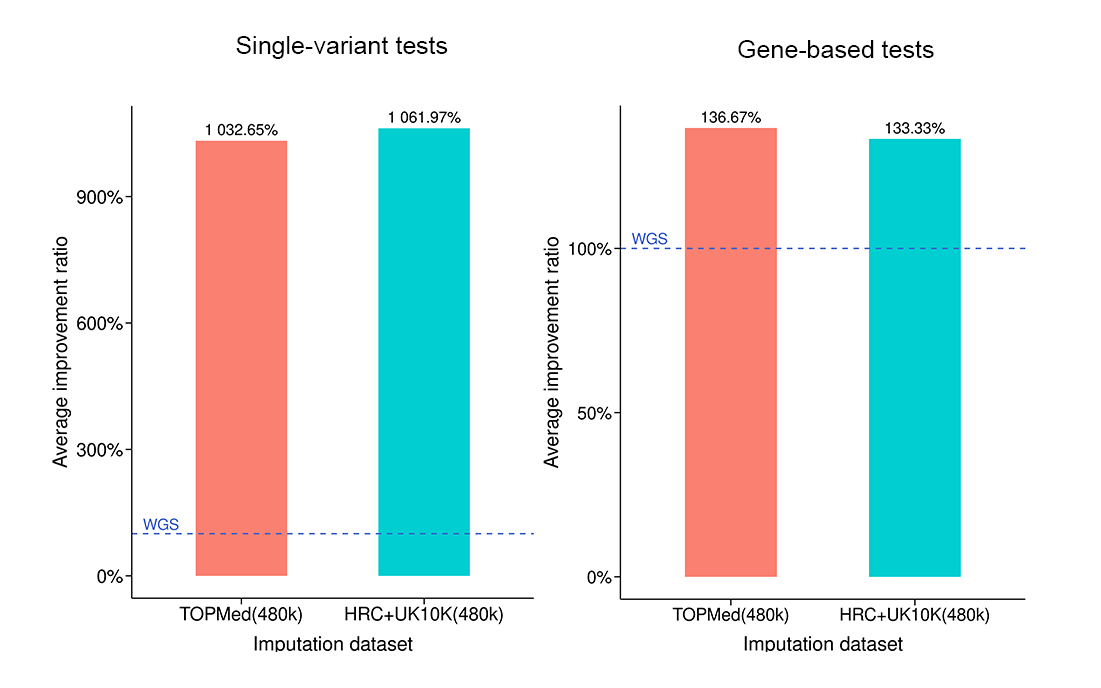

**Figure S1.** Average improvement ratio in finding significant associations in 15 complex disease association tests for imputed data (n=480 k) compared to WGS data

The ratio of TOPMed-imputed data and HUC+UK10K-imputed data were both improved by approximately 10-fold compare to WGS data in single-variant tests; For gene-based tests, the situation was in consistent with the single-variant tests but lower improvement ratio.

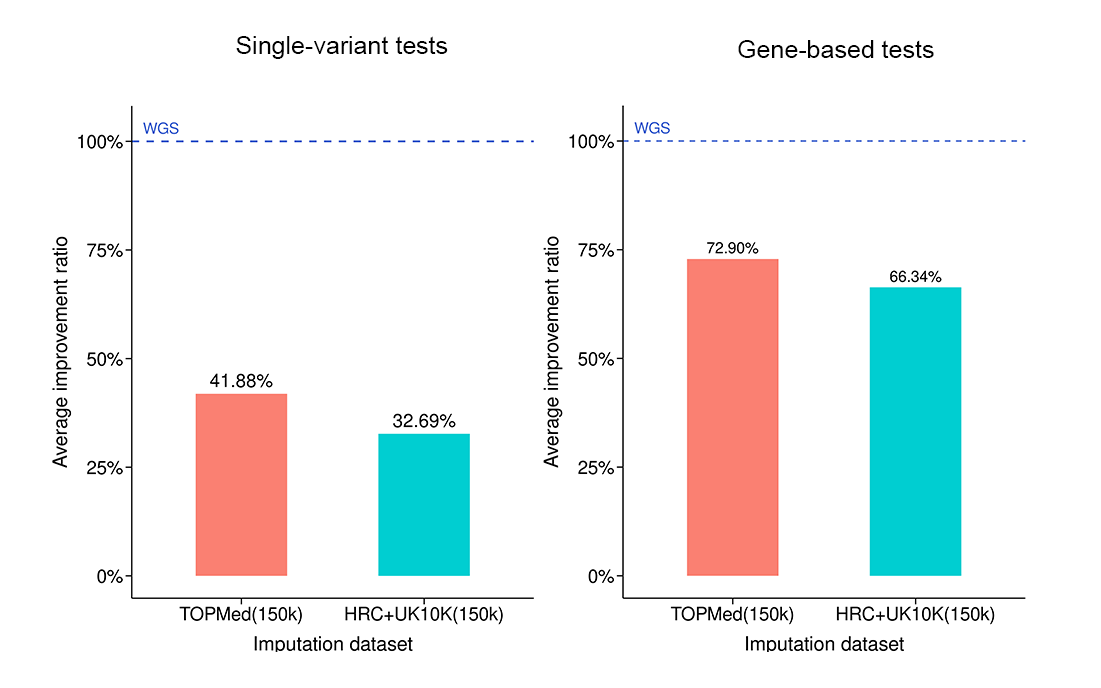

**Figure S2.** Average improvement ratio in finding significant associations in 30 biochemistry markers association tests for imputed data (n=150 k) compared to WGS data

The ratio of TOPMed-imputed data and HUC+UK10K-imputed data were both much lower than WGS data in single-variant tests ; For gene-based tests, the situation was in consistent with the single-variant tests.

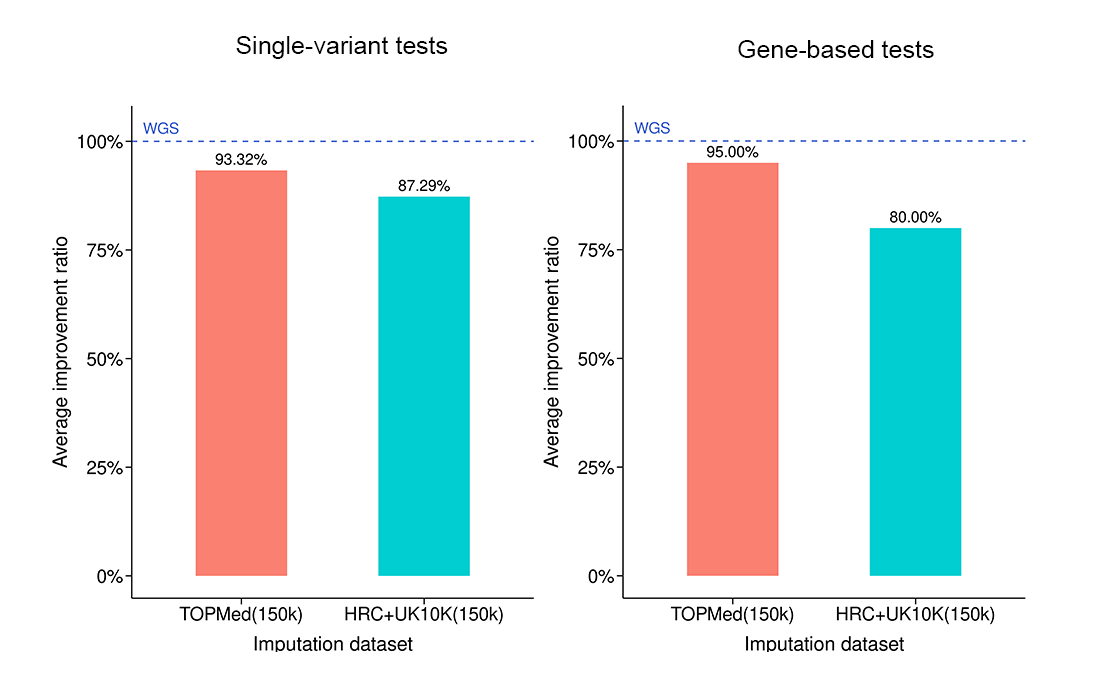

**Figure S3.** Average improvement ratio in finding significant associations in 15 complex disease association tests for imputed data (n=150 k) compared to WGS data

Although the performance of TOPMed-imputed data and HUC+UK10K-imputed data were both worse than WGS data in single-variant tests, they still closer to WGS data; For gene-based tests, the situation was in consistent with the single-variant tests and slightly better.
